# Predicting Ward Transfer Mortality with Machine Learning

**DOI:** 10.1101/2023.01.06.23284285

**Authors:** Jose L. Lezama, Gil Alterovitz, Colleen E. Jakey, Ana L. Kraus, Michael J. Kim, Andrew A. Borkowski

## Abstract

**Background:** Predicting mortality and morbidity amongst hospitalized patients has long been a struggle for inpatient Internal Medicine physicians. To prevent physician burnout, hospital organizations are turning to shift work for the care of hospitalized patients. Such shift work frequently leads to a handoff of patients, with many physicians often being the sole provider in the hospital for close to one hundred patients.

**Objectives:** We propose developing an artificial intelligence model that helps predict which patients will be most at risk of increased mortality. This model would assist providers and frontline staff in focusing their efforts on improving patient outcomes.

**Materials and Methods:** Records of patients who were transferred from non-intensive care units to intensive care units were queried from the Veteran Affairs Corporate Data Warehouse (CDW). Two thousand four hundred twenty-five records were identified. The patient outcome was designated a dependent variable, with bad outcome defined as the patient dying within 30 days of admission and good outcome as the patient being alive within 30 days of admission. Using twenty-two independent variables, we trained sixteen machine learning models, of which six best-performing ones were fine-tuned and evaluated on the testing dataset. Finally, we repeated this process with twenty independent variables, omitting the Length of Stay and Days to Intensive Care Unit Transfer variables which are unknown at the time of admission.

**Results:** The best results were obtained with the LightGBM model with both datasets, one that included Length of Stay and Days to Intensive Care Unit Transfer variables and the other without these two variables. The former achieved Receiver Operating Characteristics Curve - Area Under the Curve (ROC-AUC) of 0.89, an accuracy of 0.72, a sensitivity of 0.97, and a specificity of 0.68, while the latter achieved a ROC-AUC of 0.86, an accuracy of 0.71, sensitivity of 0.94 and specificity of 0.67 respectively.

**Conclusions:** Our predictive mortality model may offer providers a means for optimizing the utilization of resources when managing a large caseload, especially with shift changes.

## Introduction

Predicting mortality and morbidity amongst hospitalized patients has long been a struggle for inpatient Internal Medicine physicians. Sepsis, in particular, leads to over half the mortality in US Hospitals.^1^ There has been only a couple of recent models that offer some hope for an early warning system to predict worsening outcomes from sepsis.^2,3^ In order to prevent physicians from burnout, more hospital organizations are turning to shift work for the care of hospitalized patients, similar to the Internal Medicine training programs that follow the eighty-hour work limit for physicians set by the Accreditation Council for Graduate Medical Education (ACGME). Such shift work frequently leads to a handoff of patients, with many physicians often being the sole provider in the hospital for close to one hundred patients. The hand-off tools such as I-PASS and other tools can prove ineffective due to subjectivity, inconsistency of updates, and copying and pasting practices, which can negatively impact care. We propose developing an artificial intelligence model that helps predict which patients will be most at risk of increased mortality as a resource for physicians to prioritize which patients they should concentrate on, giving as much time as possible during their shift. Unfortunately, physicians tend to be reactive on shift work to calls of distress from the nursing or respiratory care or other support staff rather than proactive in addressing issues with patients before they require a transfer to an intensive care unit area. A measure called ward transfer mortality in the Veterans Healthcare System looks at the death rate of patients within 30 days of being transferred to an intensive care unit. This measure is particularly important in patients with aggressive infections such as sepsis, pneumonia, bacteremia, abscesses, and endocarditis, which require timely interventions for improved outcomes.

In our model, we attempted to look at patients who were transferred to the intensive care unit at the James A. Haley Veterans’ Hospital, a facility in the VA Health System with high complexity. This complex facility follows the usual academic center model of having trainees such as residents and fellows working with Attending staff. We examined patients with significant infectious states and those with pulmonary, neurological, and cardiology diagnoses. In addition, we looked at the following factors concerning their ward transfer mortality:

a. Drop in Leukocyte count of 50 percent or greater before they were deemed to require transfer to the intensive care unit (ICU);
b. Drop in Hemoglobin level of 25 percent or greater before they were deemed to need transfer to the ICU;
c. Presence of a C-Reactive Protein (CRP) being ordered on the patient during the hospitalization before they were deemed to require transfer to the ICU;
d. Age of the patient;
e. Prior hospitalizations before the index hospitalization were there was a transfer to the ICU.

The goal is to develop a “risk score” that would be updated at the time of any new laboratory data. Based on the risk score of those hospitalized patients, an automated patient list would be generated and prioritized from the highest risk to the lowest risk patients. This would help physicians working on these non-traditional hours shifts to prioritize those patients that possibly need further care during their shift. For example, this might lead to the following actions below depending on the clinical situation:

1. Increase use of imaging in patients with infection with source not confirmed and worsening artificial intelligence (AI) risk score;
2. Increase use of broad-spectrum antibiotics in patients with worsening AI risk score or consultation to Infectious Diseases for review of the case to ensure that proper antibiotic coverage is present and for expertise in other matters such as concomitant fungal infection in a patient with bacteremia;
3. Increase the use of advanced care settings such as Progressive Care Units (PCU) or Step-down units to attempt to manage the patient better while still avoiding intensive care unit admission;
4. Consider other advanced procedures such as bronchoscopy, transesophageal echocardiograms, or incision/debridement interventions.

A dynamic AI “risk score” model would also help prevent certain heuristic errors that can occur in clinical decision-making, such as anchoring bias.^4^

## Materials and Methods

### Dataset

Records of patients who were transferred from non-intensive care units to intensive care units were queried from the Veteran Affairs Corporate Data Warehouse (CDW). Two thousand four hundred twenty-five records were identified. The dataset did not contain any patient-identifiable information. The patient outcome was designated a dependent variable, with bad outcome defined as the patient dying within 30 days of admission and good outcome as the patient being alive within 30 days of admission. For a complete list of variables, see Table 1.

**Table 1.** List of Variables

| Variable | Description |
| --- | --- |
| Outcome | 0 = Good, 1 = Bad (death within 30 days of admission) |
| Age | Age of the patient at the time of admission |
| ED Visit Flag | Was the patient seen first in the Emergency Room |
| Admit Specialty | Admitting specialty name |
| Transfer Ward | Intensive Care Unit (ICU) Type |
| Diagnosis MDC | Diagnostic Category |
| Infection MDC | Presence of infection |
| Platelet 25 Drop Flag | 25% platelet drop |
| Lymphocytes 50 Drop Flag | 50% lymphocytes drop |
| Platelet Labs Found | Were platelet results present |
| Platelet First Lab Value | Platelet values at admission |
| Platelet Next Lab Value | Platelet values right after transfer to ICU |
| CRP Labs Found | Were CRP results present |
| Albumin Labs Found | Were Albumin results present |
| Lymphocytes First Lab Value | Lymphocytes values at admission |
| Lymphocytes Next Lab Value | Lymphocytes values right after transfer to ICU |
| Hemoglobin Labs Found | Were hemoglobin results present |
| Hemoglobin First Lab Value | Hemoglobin results at admission |
| Hemoglobin Next Lab Value | Hemoglobin results right after transfer to ICU |
| Admissions within Prev Year | Number of admissions in 12 months prior to the current admission |
| Admit to ICU Transfer Days | Days of stay from admission to ICU transfer |
| Total LOS | Total length of stay in the hospital |

The veteran population is predominantly male, which can cause the ML models not to generalize well to female veterans.^5^ However, due to the limited dataset (less than twenty-five hundred data points), we did not address this potential problem in this pilot study. Instead, we plan to address it in a follow-up project with a vastly enlarged dataset.

### Data cleaning and preparation

Columns with more than 20% missing values were deleted. Other missing values were replaced with “other” for categorical variables and with the mean value of the column for continuous variables. In addition, some numeric features were converted to categorical features with the final dataset of 8 numeric features and 14 categorical features. Finally, the data was split into training/validation (90%) and testing (10%) using the Scikit-Learn Python library.

### Data visualization

The AutoViz Python library was used to graph the dependent variable (outcome) relationship to various independent variables.

### Model training and evaluation

The PyCaret Python library was used to train, evaluate, and test various machine-learning models. Due to an imbalanced dataset (13% bad outcome, 87% good outcome), SMOTE (Synthetic Minority Oversampling Technique) was used to balance the training dataset. The six best-performing models were fine-tuned and evaluated on the unseen data from the testing dataset. The threshold was optimized for the best sensitivity to detect a bad outcome. The Light Gradient Boosting Machine (LightGBM) model showed the best results, see Figure 1 and Table 2.

**Figure 1.**
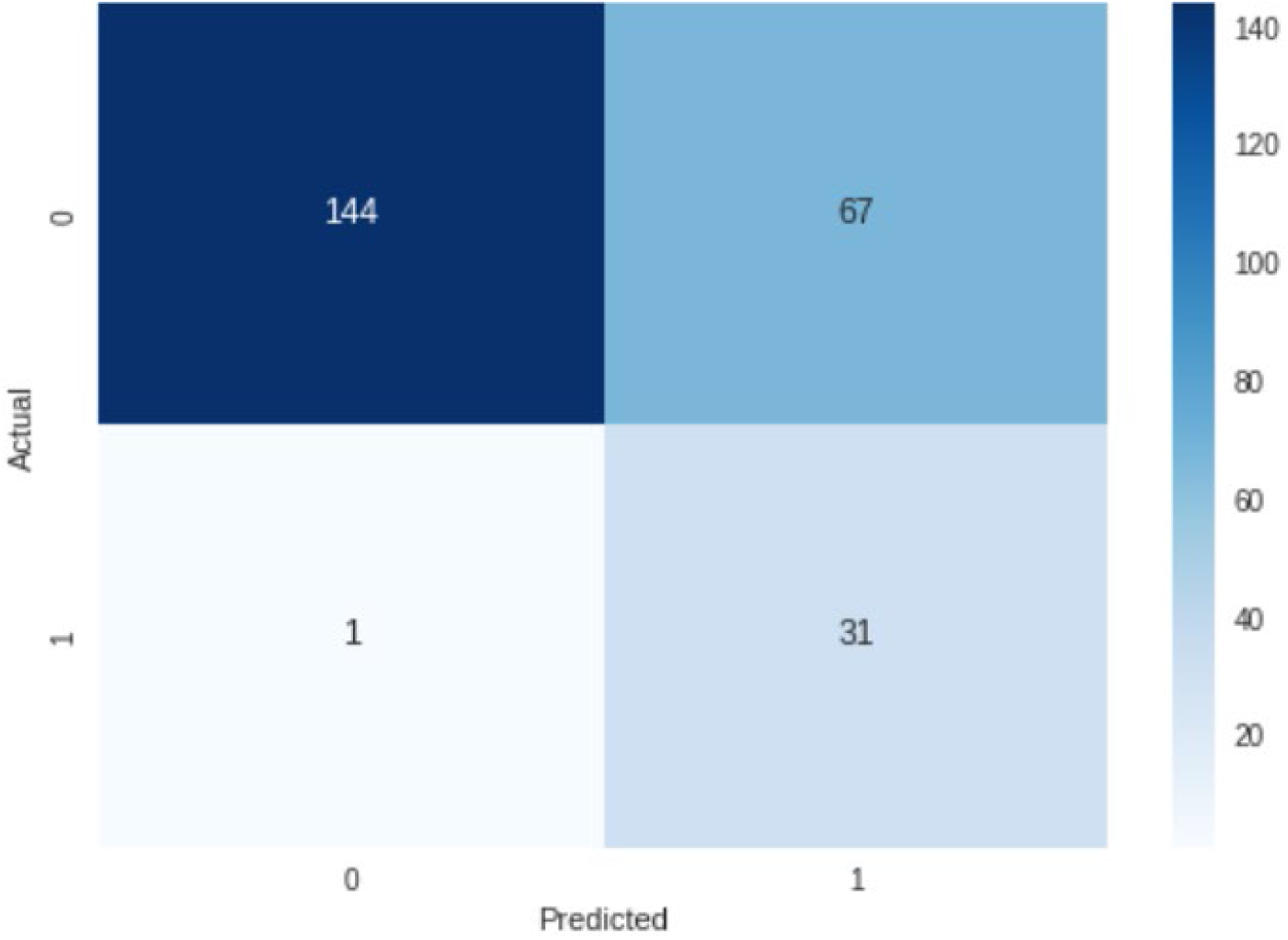
Confusion matrix for the dataset with 22 independent variables (0 – good outcome, 1 – bad outcome).

**Table 2.** Results for the dataset with 22 independent variables (lr – Logistic Regression, gbc – Gradient Boosting Classifier, lightgbm – Light Gradient Boosting Machine, qda – Quadratic Discriminant Analysis, rf – Random Forrest, xgboost – Extreme Gradient Boosting).

| ML Method | lr | GBC | lightgbm | qda | rf | xgboost |
| --- | --- | --- | --- | --- | --- | --- |
| Test Accuracy | 0.41 | 0.88 | 0.72 | 0.76 | 0.58 | 0.78 |
| Test AUC | 0.87 | 0.87 | 0.89 | 0.82 | 0.84 | 0.89 |
| Test Sensitivity | 0.97 | 0.31 | 0.97 | 0.87 | 0.94 | 0.91 |
| Test Specificity | 0.33 | 0.97 | 0.68 | 0.74 | 0.52 | 0.76 |
| Threshold | 0.1 | 0.5 | 0.1 | 0.4 | 0.2 | 0.1 |

Since two independent variables, Length of Stay and Days to Intensive Care Unit Transfer were unknown at the time of admission, we removed them from the dataset and repeated the experiments. Again, the best performance was achieved with the LightGBM. See Figure 2 and Table 3.

**Figure 2.**
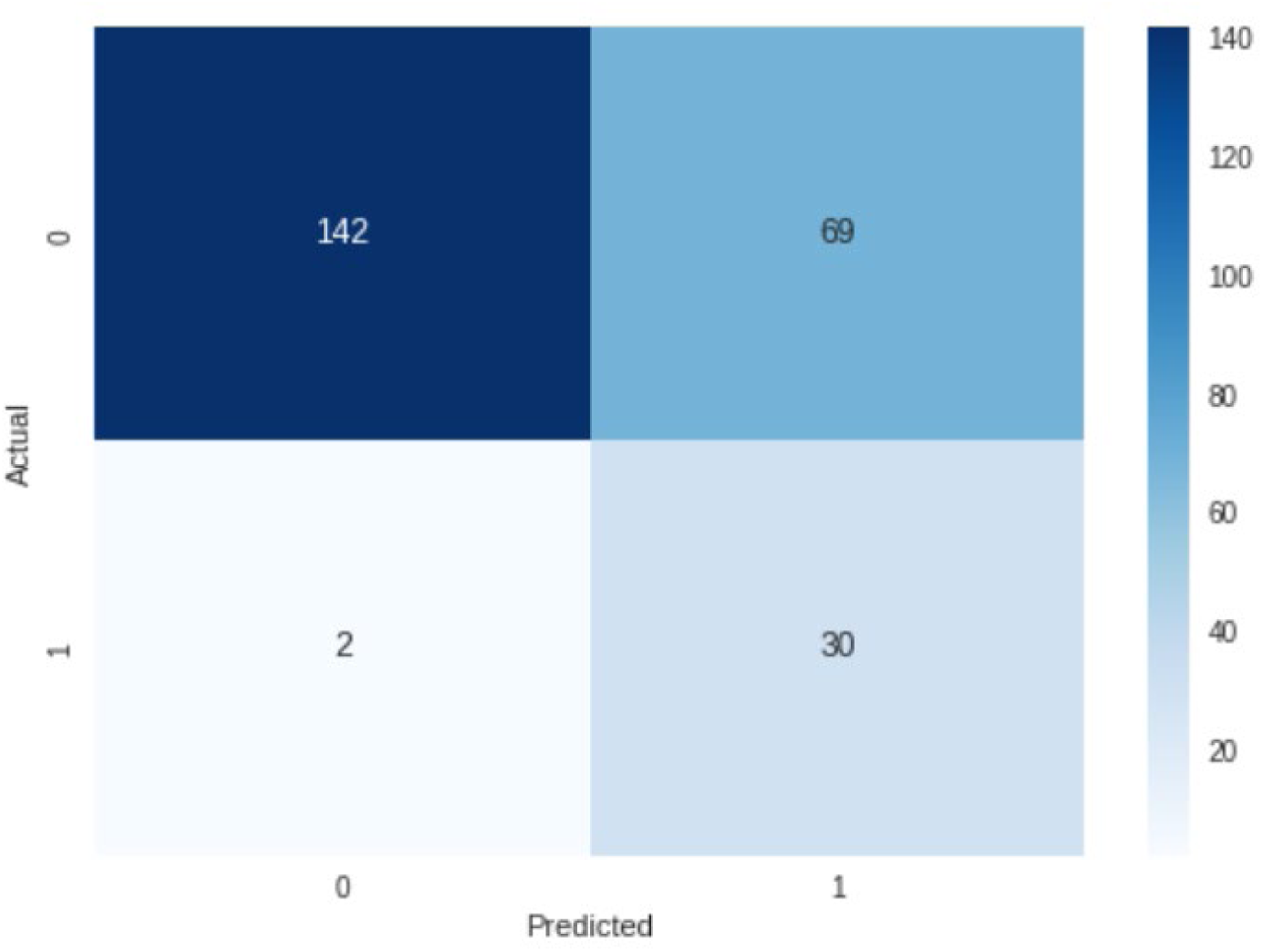
Confusion matrix for the dataset with 20 independent variables (0 – good outcome, 1 – bad outcome).

**Table 3.** Results for the dataset with 20 independent variables (without Total Length of Stay and Days to Intensive Unit Transfer).

| ML Method | lr | GBC | lightgbm | qda | rf | xgboost |
| --- | --- | --- | --- | --- | --- | --- |
| Test Accuracy | 0.37 | 0.86 | 0.71 | 0.68 | 0.62 | 0.81 |
| Test AUC | 0.83 | 0.84 | 0.86 | 0.86 | 0.83 | 0.83 |
| Test Sensitivity | 1 | 0.22 | 0.94 | 0.91 | 0.94 | 0.41 |
| Test Specificity | 0.28 | 0.96 | 0.67 | 0.65 | 0.57 | 0.87 |
| Threshold | 0.1 | 0.5 | 0.3 | 0.4 | 0.2 | 0.5 |

## Results

The best results were obtained with the LightGBM model with both datasets, one that included Length of Stay and Days to Intensive Care Unit Transfer variables and the other without these two variables. The former achieved Receiver Operating Characteristics Curve - Area Under the Curve (ROC-AUC) of 0.89, an accuracy of 0.72, a sensitivity of 0.97, and a specificity of 0.68, while the latter achieved a ROC-AUC of 0.86, an accuracy of 0.71, sensitivity of 0.94 and specificity of 0.67, on the unseen testing dataset, see Table 3 and Table 4. The top five features for the former model were Total Length of Stay, Days to Intensive Care Transfer, Lymphocytes Next Lab Value, Lymphocytes First Lab Value, and Platelet First Value, see Figure 3. For the latter model, the top five features were Lymphocytes First Lab Value, Hemoglobin First Lab Value, Hemoglobin Next Lab Value, Platelet First Lab Value, and Lymphocytes Next Lab Value, see Figure 4.

**Figure 3.**
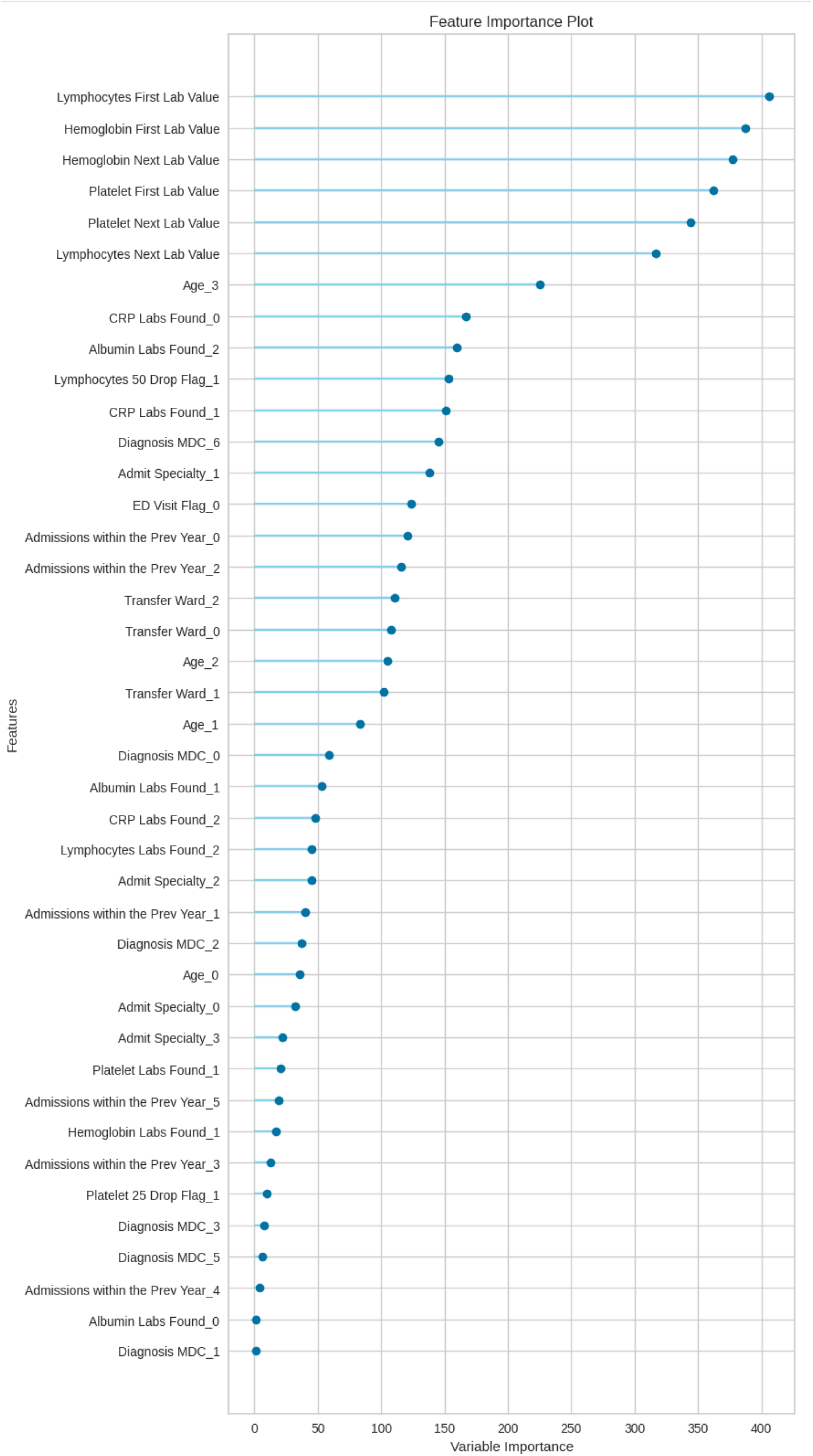
Feature importance plot for the dataset with 20 independent variables.

## Discussion

Our artificial intelligence models clearly show that there are other relationships to explore in looking at outcomes of patients that are hospitalized in the general medical wards or progressive care units (also known as step-down units) who start to have a change in lymphocyte count values, hemoglobin values, and platelet count values. Previous models looking at poor outcomes were based on length of stay models and being transferred to the intensive care unit itself.^6^ Unfortunately, these models do not help physicians proactively manage patients and recognize significant deterioration of clinical conditions before a transfer is required to an intensive care unit. Our artificial intelligence project sets out to do just that.

There are numerous future directions that we can explore as far as the expansion of this project. One direction would be to make sure that the hemoglobin drop and platelet count drop are not occurring in the setting of overt disseminated intravascular coagulation (DIC). Our project added the lymphocyte count drop, which is not part of the DIC process and related very consistently with the hemoglobin drop and the platelet count drop. The lymphocyte count, in particular, had become an essential acute phase reactant item in the face of the COVID-19 pandemic, where such dramatic lymphocyte count drops were noted when the patient started to become dramatically ill.^7^

The future direction of this project is to continue to build on additional factors that refine the predictability model of worsening of the patient’s condition once admitted to a general medical or surgical floor. For example, future projects could look at the relationship between vital signs changes in heart rate and systolic blood pressure readings, especially if they remain in what would be considered normal range but still changed from the patient’s baseline values during the initial parts of the hospitalization. In addition, future studies may also include variables like temperature, blood pressure, ventilation, ICD9/10 codes, and vasopressor data, among others.

Developing an item for physicians to use to identify patients objectively from their laboratory values changes in hemoglobin, platelet count, and lymphocyte count would be a beneficial adjunct for medical and surgical house officers who take care of numerous patients on shift work and often do not have a direct evaluation of the patient at the beginning of their shifts or even during the duration of their shifts. In addition, such an “alert” system would help stratify patients at risk of highest mortality and morbidity in the next 24 hours and also could relay those patients that have changed the most since the last time an “at risk” report was run using the variables that we have implemented in our study.

As hospitals develop teams of infectious diseases physicians and intensivists to help create high-reliability organizational states, a model like the one we have brought forward in this paper could push required targeted reviews of patients by such infectious disease physicians and intensivists. This would include such interventions as (a) vital signs being run at a much higher frequency than every shift, (b) vital orthostatic signs being obtained on such at-risk patients, (c) employing therapeutic drug monitoring on patients receiving antibiotic therapy to ensure that drug levels are reaching expected levels, (d) consideration for expedited imaging orders depending on the patient clinical situation, and (e) consideration in academic hospital models of review of patients by attending physicians if the medical house officer is a resident in training.

## Conclusions

Our pilot study created the AI predictive mortality model prototype that assists providers and frontline staff with managing large patient loads during shift change by helping them focus their efforts toward improving patient outcomes. However, further studies with more variables and a larger, more representative dataset are needed to make such a model more robust and to generalize better to various diverse populations.

## Data Availability

All data produced in the present study are available upon reasonable request to the authors

## Acknowledgments

This material is the result of work supported by the resources and the use of facilities at the James A. Haley Veterans’ Hospital. The James A. Haley Veterans’ Hospital Research and Development Committee reviewed and approved this study.

We would like to thank Mr. Scott A. Dillon for his assistance in acquiring the dataset for this study.

